# Dental teachers’ perspectives on Extended Reality in dental education: an international survey

**DOI:** 10.64898/2026.03.05.26347677

**Authors:** Ruza Bjelovucic, Bruna Neves de Freitas, Sven Erik Nørholt, Pankaj Taneja, Mette Terp Høybye, Ruben Pauwels

**Affiliations:** Department of Dentistry and Oral Health, Aarhus University, Aarhus, Denmark; Aarhus Institute of Advanced Studies, Aarhus University, Aarhus, Denmark; Department of Oral and Maxillofacial Surgery, Aarhus University Hospital, Aarhus, Denmark; Sydney Dental Hospital, Sydney, New South Wales, Australia; Sydney Dental School, University of Sydney, Australia; Interacting Minds Centre, Department of Clinical Medicine, Aarhus University; University Research Clinic for Interdisciplinary Orthopedic Pathways (UCOP), Silkeborg Regional Hospital

**Keywords:** Educational Virtual Realities, Augmented Reality, Dental Education, Attitude, Simulation Training

## Abstract

**Introduction:** Digital technologies are reshaping how health professionals are trained, and extended reality (XR) has gained attention as a tool for skills development in dental education. Yet, successful integration depends largely on educators’ perceptions, readiness, and working conditions. This study aimed to explore dental educators’ views of the educational value of XR, what barriers they experience, and how familiarity with immersive technologies relates to their use in teaching.

**Materials and Methods:** A cross-sectional, web-based survey was conducted among dental educators. The questionnaire included items on demographics, familiarity and frequency of XR use, and perceptions of educational value, barriers, and curricular integration. Descriptive statistics were calculated, and Spearman correlation analyses were performed to explore associations between familiarity, use, and perceived benefits of XR.

**Results:** Respondents reported positive attitudes toward XR, particularly for improving students’ understanding of complex anatomy (mean = 6.02/7), skill development (5.68/7), and confidence and preparedness for clinical practice (5.08-5.20/7). XR was mainly viewed as a complement to traditional teaching rather than a replacement (mean = 3.77/7). Strong correlations were observed between perceived improvements in confidence, skills, and clinical readiness (r = 0.71 - 0.89, P < 0.0001). High costs, limited technical support, and time constraints were the most prominent barriers to usage.

**Conclusion:** Overall, dental educators appear open to XR but constrained by structural and organizational factors rather than a lack of interest. Faculty development, hands-on training opportunities, and institutional support may therefore be essential to translating positive perceptions into meaningful and sustained integration of immersive technologies in dental curricula.

## 1 Introduction

Daily life has become progressively mediated by digital technologies, with various types of display systems now embedded in how people communicate, access information, and learn [1]. This reliance on digital interaction expanded during the COVID-19 pandemic, exposing limitations in conventional education and stimulating interest in more immersive approaches [2]. Extended reality (XR), an umbrella term covering virtual reality (VR), augmented reality (AR), and mixed reality (MR), has emerged as one of such directions. XR offers applications such as embodied learning, where users appear as avatars and engage with others in shared virtual environments [3, 4]. Despite technological developments in the past decade, the educational sector has been slow to integrate contemporary technological advances into daily education [5].

These dynamics are similarly reflected in dental education, which is also confronted with the need for curricular reform [6]. Many dental programs continue to face structural constraints, including limited access to real patients, insufficient infrastructure, and variation in competency standards across institutions [7]. These conditions contribute to persistent discrepancies between students’ self-assessed readiness for independent practice and faculty evaluations of their clinical readiness [8].

Within dental education, XR technologies are often praised for enabling self-paced learning [9, 10], unlimited repetition [9], providing immediate feedback [11], and facilitating the acquisition of psychomotor and procedural skills [12, 13]. They offer a low-risk environment in which students can practise repeatedly without compromising patient safety [14, 15]. Several studies suggest that such immersion may contribute to reduced anxiety and greater perceived preparedness for clinical training [16, 17]. XR-based learning has also been associated with gains in theoretical knowledge and self-confidence when compared with traditional instructional methods [14]. Moreover, most research reports generally positive student attitudes toward the use of XR in dental curricula.

However, the successful integration of XR in education also depends on educators’ decisions, beliefs, and competencies [18]. Existing literature on dental faculty perspectives tends to be limited to single-site studies or focuses on only one aspect of XR, such as virtual or augmented reality. Across dental, medical, and surgical education, reports indicate a positive attitude toward XR modalities among teaching staff [19–21]. Broader acceptance of XR and comprehensive insights into dental educators’ perspectives remain limited.

Therefore, this study aims to provide an overview of the current landscape of challenges, barriers, and educators’ expectations for implementing XR in dental education. The following research questions drive this study:

1. What is the dental educator’s perceived educational value of XR technology?
2. What barriers do dental educators perceive as limiting the integration of extended reality technologies into teaching practice?
3. How familiar are dental educators with XR technologies, and how does familiarity relate to attitudes toward curricular integration of XR?
4. Is educators’ use and experience with immersive technologies associated with perceived teaching benefits of XR?
5. Do dental educators perceive XR as a replacement for, or a complement to, traditional teaching methods?

## 2 Materials and methods

This study was designed as a cross-sectional web-based survey. Ethical approval was obtained from the Aarhus University Research Ethics Committee (Approval 2025-012). The CHERRIES guidelines were followed in tailoring this survey [22]. Participation was voluntary and anonymous, and informed consent was obtained electronically before respondents accessed the questionnaire.

The target population were educators within dental education. Inclusion criteria were dental and health professionals actively involved in the education of dental students, regardless of academic position or clinical teaching experience. A combination of convenience and snowball sampling was used. Initially, email invitations were distributed to educators listed in the Aarhus University teaching database. To reach globally, professional mailing lists, social media, and academic networks were used. Participants were also encouraged to share the survey with eligible colleagues within their institutions and professional communities. Potential survey participants received email invitations in early September 2025. The survey was open to completion until late November 2025.

The survey (see Appendix A) was adapted from the pre-existing, validated survey developed by Khukalenk et al. [23] and Wozney et al. [24], which examines attitudes toward educational technologies in the classroom. Relevant Likert-scale items from these tools were modified to reflect the specific context of dental teaching and XR integration. To optimize completion rates and respect respondents’ time, the survey was condensed to 26 questions. XR specialists, experienced dental educators, and an experienced teacher in medical anthropology reviewed the items to ensure the survey’s content validity.

Consequently, the final survey consisted of 26 questions divided into the following three sections:

- Section 1: General information: gender, age, teaching experience, subjects taught, and the country where the university is located.
- Section 2: Familiarity and frequency of usage of XR (VR, MR, and AR) in teaching, using a Likert scale.
- Section 3: Perceptions of XR in teaching and integration level, containing 19 Likert-scale questions using a consistent seven-point scale: 1-Strongly Disagree, 2-Disagree, 3-Slightly Disagree, 4-Neutral, 5-Slightly Agree, 6-Agree, and 7-Strongly Agree. This section has four subgroups of questions focusing on: a) educational values of XR, b) impact on teaching practice, c) barriers to implementation, and d) attitudes, concerns, and curriculum integration.

The survey was administered in English via the REDCap platform with an estimated completion time of approximately eight minutes.

### 2.1 Data Analysis

All data were analysed using GraphPad Prism (version 10.6.1 for Windows, GraphPad Software, Boston, MA, USA). Quantitative data were examined using descriptive statistics, including means and standard deviations. Spearman correlation coefficients (R) were calculated to explore associations among familiarity with XR, frequency of XR use, and educators’ perceptions. A significance level of 0.05 was used with a Holm-Bonferroni correction for multiple testing. For the reader’s convenience, adjusted p-values will be shown throughout the results. To check the reliability of the adapted questionnaire, the Standardized Cronbach’s alpha (α) was calculated.

## 2 Results

The adapted instrument demonstrated good internal consistency (Cronbach’s α = 0.843).

### 3.1 Respondent demographics

A total of 41 survey responses were collected from 15 countries (Australia, Belgium, Denmark, Egypt, France, Hong Kong, India, Ireland, Norway, Portugal, Romania, Sweden, Switzerland, The Netherlands, USA). One response was excluded due to incomplete data, resulting in 40 valid responses for analysis. Some participants left one question unanswered but were still included in the statistical analysis. Therefore, the total number of responses for certain questions was 38 or 39. Of the participants, 46.3% were female, and 53.7% were male. The most represented age group was 45-54 years (41.5%), followed by 35-44 years (26.8%), 25-34 years (14.6%), and 55-64 years (9.8%). Consistent with this distribution, the largest proportion of respondents reported more than 20 years of teaching experience (29.3%), followed by those with 1-5 years of teaching experience (26.8%). Participants teach a range of dental specialties, with the highest proportions coming from oral radiology, endodontics, periodontology, and restorative dentistry, respectively (Figure 1).

**Figure 1.**
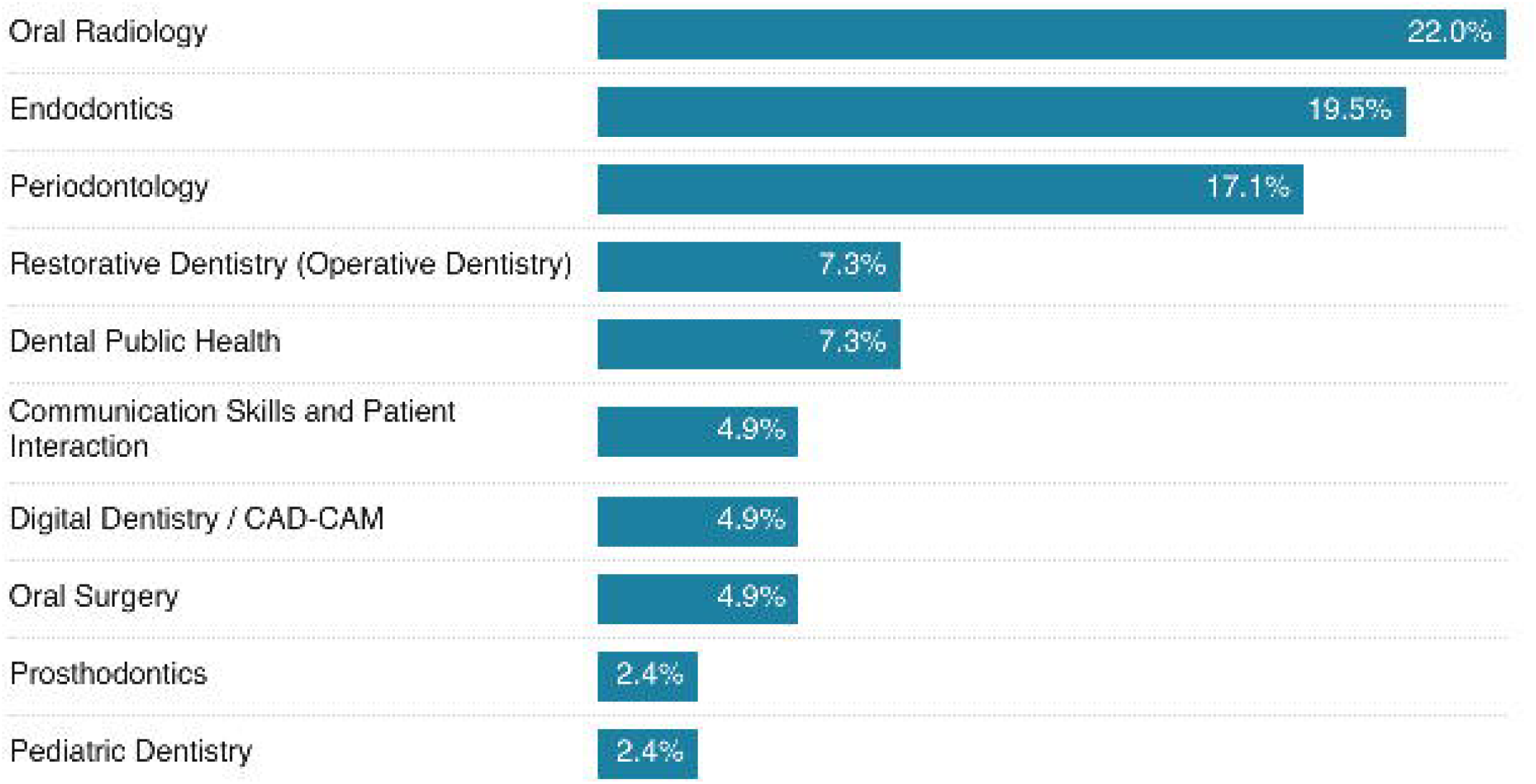
Field of teaching of respondents to the survey

**Figure 2.**
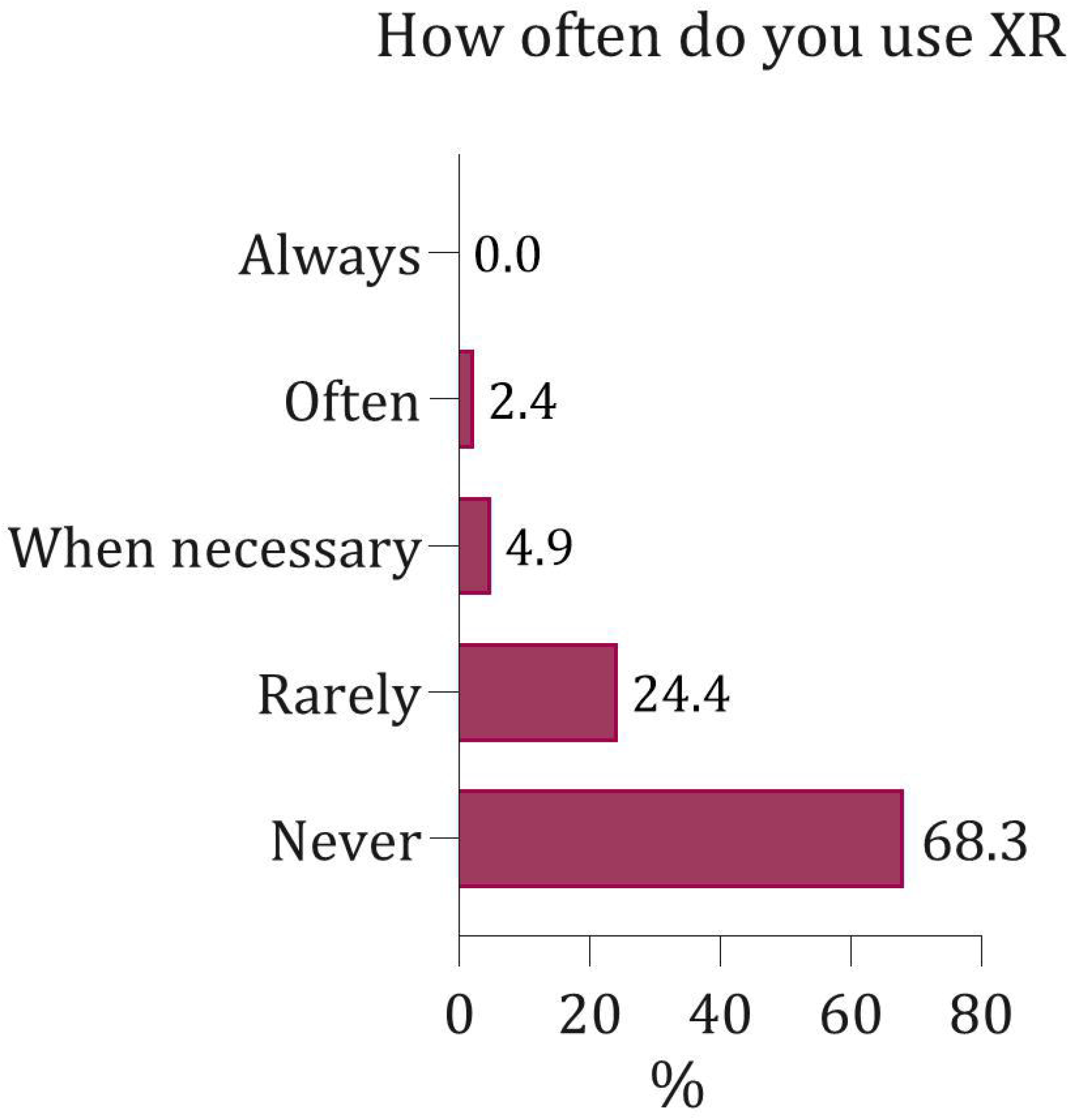
Frequency of use of XR in teaching

### 3.2 Questionnaire analysis

Descriptive statistics for all 19 Likert-scale statements are presented in Table 1. Seventeen statements had a mean score above four (neutral midpoint), indicating an overall positive perception of XR in dental education. Only two statements (5, 11) yielded mean scores below the neutral midpoint, indicating lower agreement. The strongest agreement was observed for the statement *“Immersive technology improves students’ understanding of complex anatomical structures*, with 56.1% of respondents indicating agreement and a mean score of 6.02 (SD = 1.01).

**Table 1.**
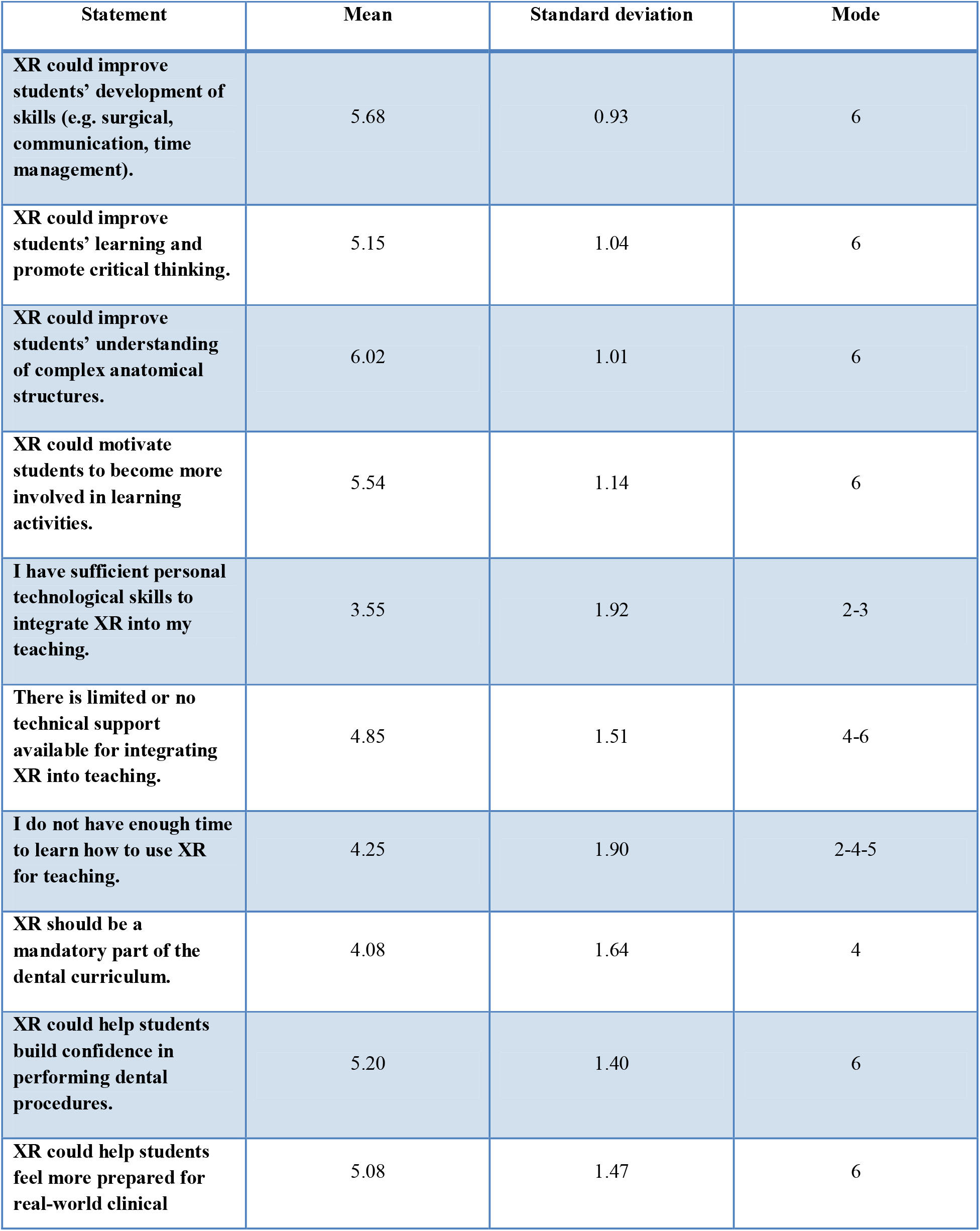
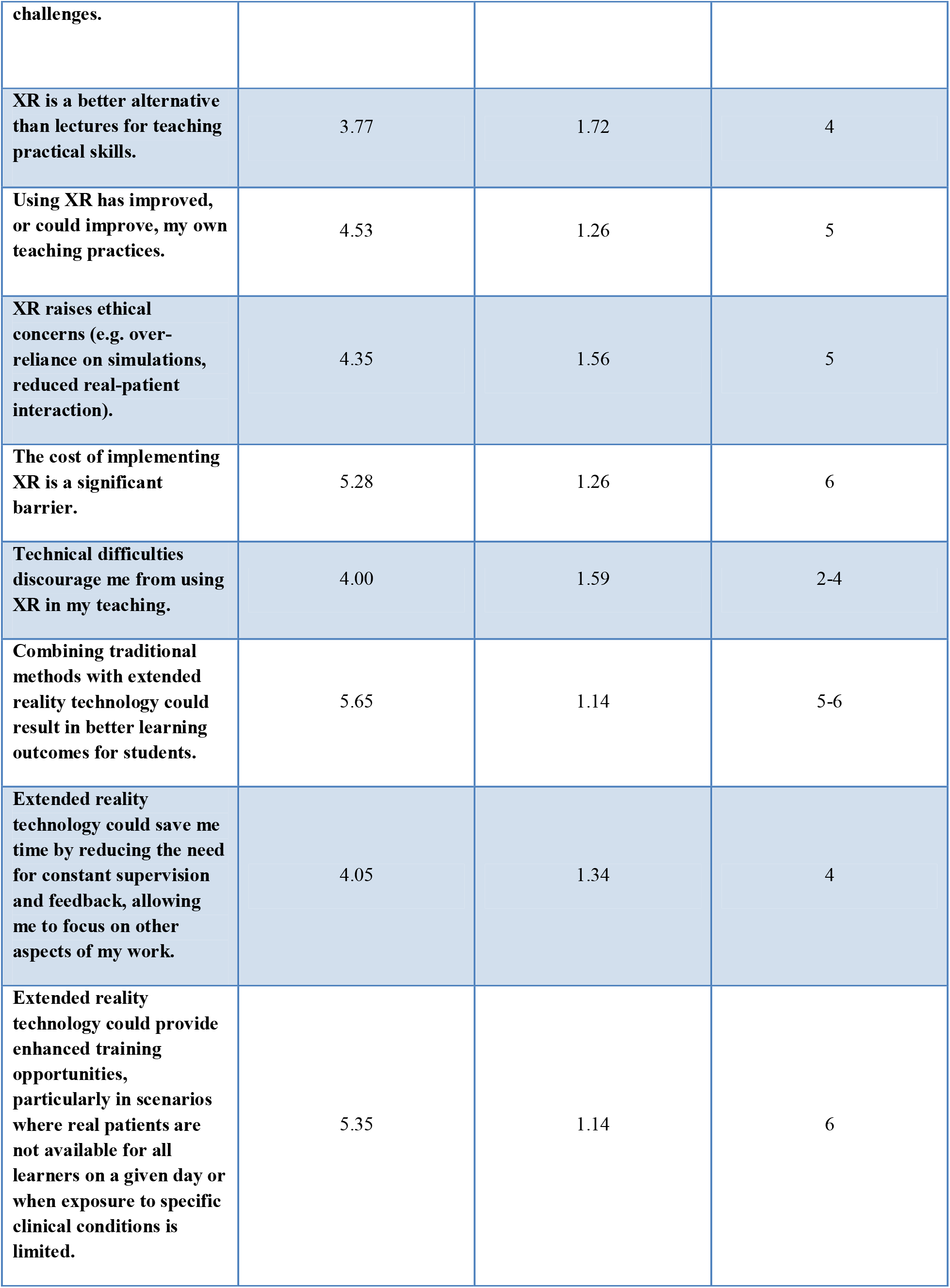
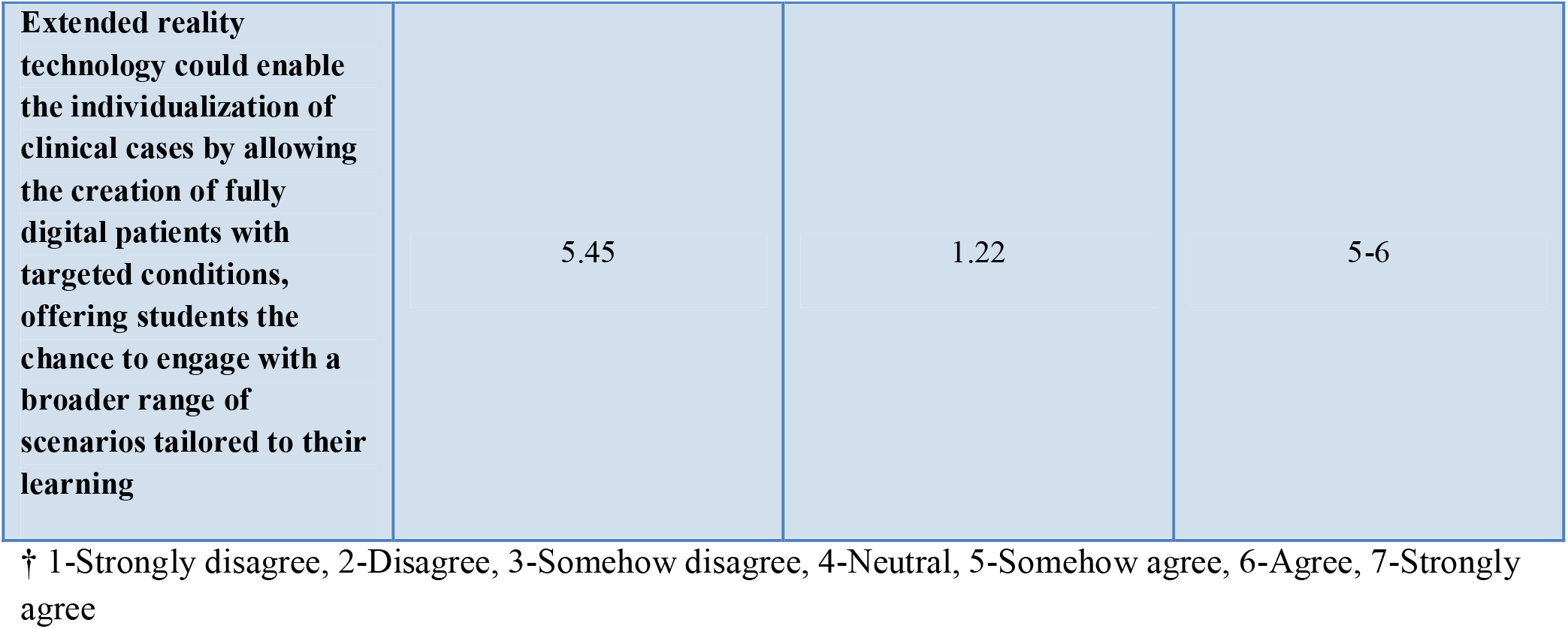
Dental teachers’ perceptions towards Extended Reality (XR) in dental education.

#### 3.2.1 Research question 1: What is the teacher’s perceived educational value of XR technology

Seven Likert-scale items assessed dental teachers’ perception toward XR technology in dental education (1, 2, 3, 4, 9, 10, 16). The majority of respondents expressed moderately positive perceptions across this domain. Dental educators generally agreed that XR technology could enhance students’ skill development, promote critical thinking, increase motivation for learning, and support confidence building and preparedness for real-world clinical challenges. Further analysis indicated a high level of agreement that combining immersive technology with traditional teaching approaches may lead to improved learning outcomes (mean = 5.65, SD = 1.14). Respondents generally saw XR as useful for training during patient cancellations or limited clinical exposure (mean = 5.35, SD = 1.14). There was a strong positive association between the perception that XR could help students build confidence and the perception that XR could help students feel more prepared for real-world clinical challenges (R = 0.88, p < 0.0001). Another strong positive association was observed between the perception that XR could enable the individualization of clinical cases and the perception that XR could enhance training opportunities (R = 0.84, p < 0.0001). A positive association was also identified between the perception that XR could enable individualization and the perception that XR could improve students’ skill development (R = 0.71, p < 0.001), as well as between the perception that XR could improve students’ skill development and their understanding of complex anatomical structures (R = 0.71, p < 0.0001).

#### 3.2.2 Research question 2: What barriers do dental educators perceive as limiting the integration of extended reality technologies into teaching practice?

Seven questions explored barriers that may limit the integration of XR into teaching practice (5, 6, 7,14, 15, 17). The most prominent barrier was the cost of implementing XR, which received a mean score of 5.28 ± 1.26. A perceived lack of technical support was also commonly reported (mean = 4.85, SD = 1.51). Respondents indicated time constraints as a barrier, with mixed responses regarding whether they had sufficient time to learn how to use XR for teaching purposes (mean = 4.25, SD = 1.90). Moreover, respondents expressed overall general agreement regarding whether XR could save time by reducing supervision and allowing more focus on other aspects of their work (mean = 4.05, SD = 1.34). In addition, participants had mixed views on whether technical difficulties discourage them from using XR (mean = 4.00, SD = 1.59). Self-reported technological competence was comparatively low, with respondents tending to disagree that they have sufficient personal technological skills to integrate XR into their teaching (mean = 3.55, SD = 1.92).

#### 3.2.3 Research question 3: How familiar are dental educators with XR technologies, and how does familiarity relate to attitudes toward curricular integration of XR?

Overall, respondents reported a low to moderate level of familiarity with XR technologies (Table 2). Familiarity differed across modalities, with the highest reported for VR, followed by AR, and the lowest reported for MR. A statistically significant gender difference was observed for AR familiarity, with male educators reporting higher familiarity than female educators (p = 0.035) (Table 2). XR technologies were infrequently used in teaching (Figure 3). More than two-thirds of respondents reported never using XR, 24.4% reported rare use, and 4.9% indicated using XR when necessary.

**Table 2.** Average level of familiarity of respondents with different XR modalities, per gender.

| **Familiarity with** | **Female** | **Male** | **P-value** |
| --- | --- | --- | --- |
| **VR** | 2.43 | 2.86 | 0.157 |
| **AR** | 1.95 | 2.86 | 0.035 |
| **MR** | 1.90 | 2.36 | 0.282 |

† VR-virtual reality, AR-augmented reality, MR-mixed reality. 1-not familiar, 2- slightly familiar, 3- somewhat familiar, 4- familiar, 5- very familiar

#### 3.2.4 Research question 4: Is educators’ use and experience with immersive technologies associated with perceived teaching benefits of XR?

No significant association was found between the educators’ use and experience with immersive technologies associated with perceived teaching benefits of XR. When asked whether XR could improve their teaching practices, respondents expressed overall neutral views (mean = 4.53, SD = 1.26).

#### 3.2.5 Research question 5: Do dental educators perceive XR as a replacement for, or a complement to, traditional teaching methods?

On average, respondents expressed a divided view regarding whether XR should be a mandatory part of the dental curriculum (mean = 4.08, SD = 1.64). Further analysis shows that, on average, respondents tended to slightly disagree that XR is a better alternative than lectures for teaching practical skills (mean = 3.77, SD = 1.72). Table 3 shows the distribution of responses for these statements.

**Table 3.** Respondents’ level of agreement on the statements that XR should be a mandatory part of the dental curriculum and that XR is a better alternative than lectures for teaching practical skills.

|  | **XR mandatory part of curriculum (%)** | **XR better alternative than lectures (%)** |
| --- | --- | --- |
| **Totally agree** | 10.0 | 13.2 |
| **Agree** | 7.5 | 10.5 |
| **Somehow agree** | 22.5 | 18.4 |
| **Neutral** | 25.0 | 28.9 |
| **Somehow disagree** | 15.0 | 10.5 |
| **Disagree** | 15.0 | 10.5 |
| **Totally disagree** | 5.0 | 7.9 |

† XR – Extended Reality

## 4 Discussion

The present study examined dental educators’ perceptions of XR’s role in dental education. Overall, the findings suggest that XR is a valuable pedagogical tool, particularly in skill development, confidence building, and understanding of complex anatomical structures. Despite these positive perceptions, XR was not viewed as a replacement for conventional teaching of practical skills. This is consistent with findings from systematic reviews, which indicate that VR, AR, and MR technologies are mostly viewed as supportive tools in dental education [12, 25–27]

The results showed positive correlations between educators’ self-reported familiarity and their frequency of use of immersive technologies, with clear differences across modalities (VR: R = 0.315; AR: R = 0.639; MR: R = 0.661). Contrary to expectations, overall familiarity levels were relatively low (VR: 2.65/5; AR: 2.29/5; MR: 2.10/5), indicating that many educators are at an early stage of exposure. These findings are in accordance with de Boer et al. [28], indicating that fewer than 15-20% of dental schools worldwide have implemented XR. This finding also suggests that simply knowing what VR/AR/MR is does not necessarily lead to actual use and integration in teaching. It is therefore suggested that providing hands-on training may help to move from awareness to routine educational integration, especially for AR and MR, where familiarity is closely linked to frequency of use.

Strong correlations between perceived improvements in students’ confidence and skills, preparedness for clinical practice, and opportunities for individualized training suggest that dental educators primarily view XR as a tool to enhance clinical readiness. Several factors could explain this perception. Firstly, previous research indicates that performance in VR-based simulation can help identify students who require additional training and that simulator performance may predict clinical competence [29, 30]. This predictive potential supports the development of individualized XR scenarios, where learners who struggle can receive targeted practice while more advanced students can practice a higher level of skills [15, 26]. Secondly, MR and VR simulation environments enable repetitive, self-directed practice of psychomotor skills, which has been shown to support improvements in manual dexterity [13, 31, 32]. This repeated practice may reduce the need for continuous direct supervision, allowing instructors to allocate time more efficiently while maintaining training quality [25]. Finally, the literature consistently highlights the value of XR in the early stages of skills acquisition, particularly in preclinical education [21, 25, 32, 33]. Immersive simulation provides a low-risk environment in which students can practice procedures, learn from errors, and build confidence without the stress of real patient care [29]. This safe learning context may help reduce performance anxiety and facilitate the transition to clinical settings [15].

Cost, technical support, and time constraints emerged as the most prominent barriers to implementation. Collectively, these findings indicate that the current challenge in XR integration lies not in perceived educational value but in practical and organizational factors. A study investigating dental educators’ perceptions of VR in education has identified high equipment costs as one of the primary barriers to implementation, alongside limitations in available features, lack of acceptance, and generally dismissive views of the technology [21]. Similar findings, highlighting cost as a major challenge for integrating VR into the dental curriculum, have been reported [15]. Likewise, the initial implementation of MR in endodontic education has been shown to require substantial upfront investment [34]. However, when considered from a long-term perspective, VR-based training may help reduce overall educational costs by decreasing the need for students’ direct contact hours with educators [15, 21] and allowing repeatable training [29, 35, 36]. In addition, XR-based simulation has been reported to reduce costs in preclinical dental education by decreasing reliance on consumables (e.g., artificial teeth, dental materials) and lowering the maintenance demands of traditional phantom-head laboratories [37]. Financial barriers could also be overcome with collaborations with XR firms, public-private partnerships, and the use of open-source XR tools [19]. However, the overall quality of the XR must be ensured and should demonstrate effectiveness comparable to or greater than traditional methods. Regarding technical support, providing workshops and creating technical platform accounts might help achieve a higher degree of acceptance of XR [19]. Notably, educators showed moderate agreement that insufficient time limits engagement with XR. In an online survey, Bencharit et al [21] reported that 13% of respondents indicated they did not have sufficient time to learn VR or integrate it into their curriculum. The adoption of new educational technologies is time-consuming; however, improved access to structured training resources, digital learning platforms, and on-demand instructional materials may facilitate a smoother transition [21]. A weak negative correlation (R = −0.251) was observed between years of teaching and perceived time constraints. Although this trend should be interpreted cautiously, it may reflect broader differences in attitudes toward educational innovation across career stages. Previous research suggests that senior faculty may express skepticism toward XR, partly because it could disrupt traditional pedagogical approaches [19]. Concerns about the time required to learn new technologies, the potential impact on workload, and limited institutional support or scarce time for course development have previously been identified as barriers to faculty engagement in technology-enhanced education [38]. These structural factors may therefore contribute to the perception that XR adoption is difficult to accommodate within existing academic responsibilities.

There are several limitations in this study that should be acknowledged. The findings were based on self-reported questionnaire data, which may introduce response bias and affect the reliability and validity of the measurements. The generalizability of the findings may be limited by the scope and demographic characteristics of the sampled population, and the results may not fully capture the diversity of perspectives across the wider population of dental educators. The educational value of XR may differ across subject areas and educators ‘specialties, and a discipline-specific survey could provide more nuanced insights. Establishing international collaborative networks or working groups focused on XR in dental education could also facilitate in-depth qualitative studies and promote the exchange of best practices, thereby supporting more informed and coordinated implementation of XR.

## 5 Conclusion

The findings of the present study indicate that dental educators are pedagogically receptive to XR but face practical barriers related to time, cost, and technical infrastructure. The positive correlations between familiarity and frequency of use suggest that targeted faculty development, hands-on training, and institutional support structures may be critical for translating positive attitudes into successful educational integration. Overall, the findings indicate that successful integration of XR in dental education will depend on institutional support, structured training, and strategic implementation efforts.

## Acknowledgements

The authors thank all anonymous participants in this survey and those who helped with its circulation.

## Author Contributions

Conceptualization, R.B., M.T.H., and R.P.; Data curation, R.B., P.T., and R.P; Funding acquisition, R.P.; Formal analysis, R.B., B.N.F, M.T.H, and R.P; Investigation, R.B.; Methodology, R.B., B.N.F, M.T.H, and R.P.; Project administration, R.B. and R.P.; Supervision, R.P.; Writing-original draft, R.B., R.P.; Writing-review and editing, B.N.F., S.E.N., P.T., and M.T.H.

All authors have read and agreed to the published version of the manuscript.

## Data Availability Statement

Data will be available from the corresponding author on reasonable request.

